# Deciphering the consequence of deep intronic variants: a progeroid syndrome caused by a *TAPT1* mutation is revealed by combined RNA/SI-NET sequencing

**DOI:** 10.1101/2022.07.15.22276800

**Authors:** Nasrinsadat Nabavizadeh, Annkatrin Bressin, Poh Hui Chia, Ricardo Moreno Traspas, Nathalie Escande-Beillard, Carine Bonnard, Zohreh Hojati, Scott Drutman, Susanne Freier, Mohammad El-Khateeb, Rajaa Fathallah, Jean-Laurent Casanova, Wesam Soror, Alaa Arafat, Mohammad Shboul, Andreas Mayer, Bruno Reversade

## Abstract

Exome sequencing has introduced a paradigm shift for the identification of germline variations responsible for Mendelian diseases. However, non-coding regions, which make up 98% of the genome, cannot be captured. The lack of functional annotation for intronic and intergenic variants makes RNA-seq a powerful companion diagnostic. Here, we illustrate this point by identifying six patients with a recessive Osteogenesis Imperfecta (OI) and neonatal progeria syndrome. By integrating homozygosity mapping and RNA-seq, we delineated a deep intronic *TAPT1* mutation (c.1237-52G>A) that segregated with the disease. Using patients’ fibroblasts, we document that TAPT1’s nascent transcription was not affected, indicating instead that this variant leads to an alteration of pre-mRNA processing. Predicted to serve as an alternative splicing branchpoint, this mutation causes *TAPT1* exon 12 skipping, creating a protein-null allele. Additionally, our study reveals dysregulation of pathways involved in collagen and extracellular matrix biology in disease-relevant cells. Overall, our work highlights the power of transcriptomic approaches in deciphering the repercussion of non-coding variants as well as in illuminating the molecular mechanisms and underlying pathways of human diseases.

## INTRODUCTION

Whole exome sequencing (WES) targets less than 2% of our genome whereas the majority of non-coding sequences are still understudied. These sequences are crucial for gene regulation, are to a large extent transcribed and form a significant portion of our genome which are also susceptible to harbor variants responsible for human diseases ^1–4^. Indeed, from the more than 4000 Mendelian phenotypes reported to date, approximately 50% still lack the identification of the underlying genetic cause ^5^. This speaks to the necessity to further explore non-coding sequences. Whole-genome sequencing (WGS) provides a more comprehensive method to cover the full genome, however, a key challenge to its implementation is the prioritization of the vast amount of non-coding variants identified. This barrier to interpretation is in part driven by the lack of annotated information in intronic and intergenic regions which together comprise up to 98% of our genome. RNA-sequencing (RNA-seq) has proven to be a powerful complementary approach that can help overcome these challenges by revealing the functional impact of the genetic variants at the transcriptome level. The use of RNA-seq in conjunction with WGS permits cross-referencing of endogenous RNA levels and splicing events to help prioritize disease-causing mutations at the DNA level ^6–8^.

Here we report the study of six affected children from two consanguineous 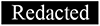 families that presented with a congenital syndrome consisting of osteogenesis imperfecta (OI), severe developmental delay and neonatal progeria. By combining homozygosity mapping, RNA-seq and targeted Sanger sequencing, we identified an intronic homozygous variant (c.1237-52 G>A) in *TAPT1* (MIM612758) which entirely segregated with the disease. Our downstream characterization assays using patient-derived fibroblasts showed how this private non-coding mutation induces the complete skipping of exon 12 leading to a TAPT1 protein-null allele.

*TAPT1* codes for a predicted transmembrane protein which has been related to ER/Golgi pathways, human Cytomegalovirus (HCMV) infection and primary ciliogenesis ^9–15^. Our functional studies using patient derived TAPT1-knockout cells could not detect patent anomalies in the pathways previously linked to TAPT1 indicating that its precise molecular function has yet to be ascertained. Notwithstanding, our RNA-seq and NET-seq (native elongating transcript sequencing) analyses revealed a role for TAPT1 in collagen and ECM biology, which is consistent with clinical presentation of our patients. Overall, our study highlights the capacity of applying robust transcriptomic approaches to prioritize disease-causing genes and understand the underlying pathogenesis of Mendelian disease.

## RESULTS

### A severe recessive progeroid syndrome with osteogenesis imperfecta

We investigated six severely-affected children from two consanguineous 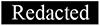 families (Fig. 1A, 1B) manifesting growth retardation, short stature, multiple bone deformities, lipodystrophy and neonatal progeria. The patients had various craniofacial abnormalities including prominent forehead, plagiocephaly, depressed nasal bridge, nasal septum deviation, low set ears, ear deformities, micrognathia, and occult cleft palate (Fig. 1C - 1E, S1). The patients also suffered from microphthalmia, cataract, and bilateral esotropia. They had translucent, wrinkled skin with patent acrogeria and sparse hair with premature depigmentation (Fig. 1C - 1E, S1). They also displayed pectus excavatum and brachydactyly of both hands and feet (Fig. 1C - 1E, S1). X-rays of patient V.1 from family 1 showed extensive deformity of the bones, bone dysplasia with bowing, and evidence of previous multiple fractures (Fig. 1F). The proband had spared joints, a flattened epiphysis of the humeral bone, irregular growth of arm bones resulting in small deformed radius bone, and a bowed ulnar bone. She also presented a deformed clavicular bone with displacement of both claviculosternal and acromioclavicular joints, deformed shoulders, irregular development of the scapula, bilateral shallow acetabulum, abnormal contour of bilateral femoral head, and absent femoral diaphysis. X-rays also revealed severe calcification defects involving premature atherosclerotic vascular calcification, periarticular soft tissue calcification, and irregular calcification of carpal bones (Fig. 1F). Brain abnormalities were also reported in patient V.1 (F1) with cranial MRI showing defects in the white matter of the frontal and occipital lobes with pachygyria, possibly representing some form of leukodystrophy. The proband V-1 (F1) and V-5 (F1) died of severe respiratory infection and inflammation at the age of 10 and 4.5 years, respectively. The history of a similar disease was remarkable in this extended kindred. Two affected girls (IV-7 (F1) and V-13 (F1)) born to the mother’s aunts who showed similar clinical manifestation and died of severe respiratory distress at the age of 5 years. Another case (V-12 (F1)) of 2 years of age is alive and manifests similar clinical features.

**Figure 1.**
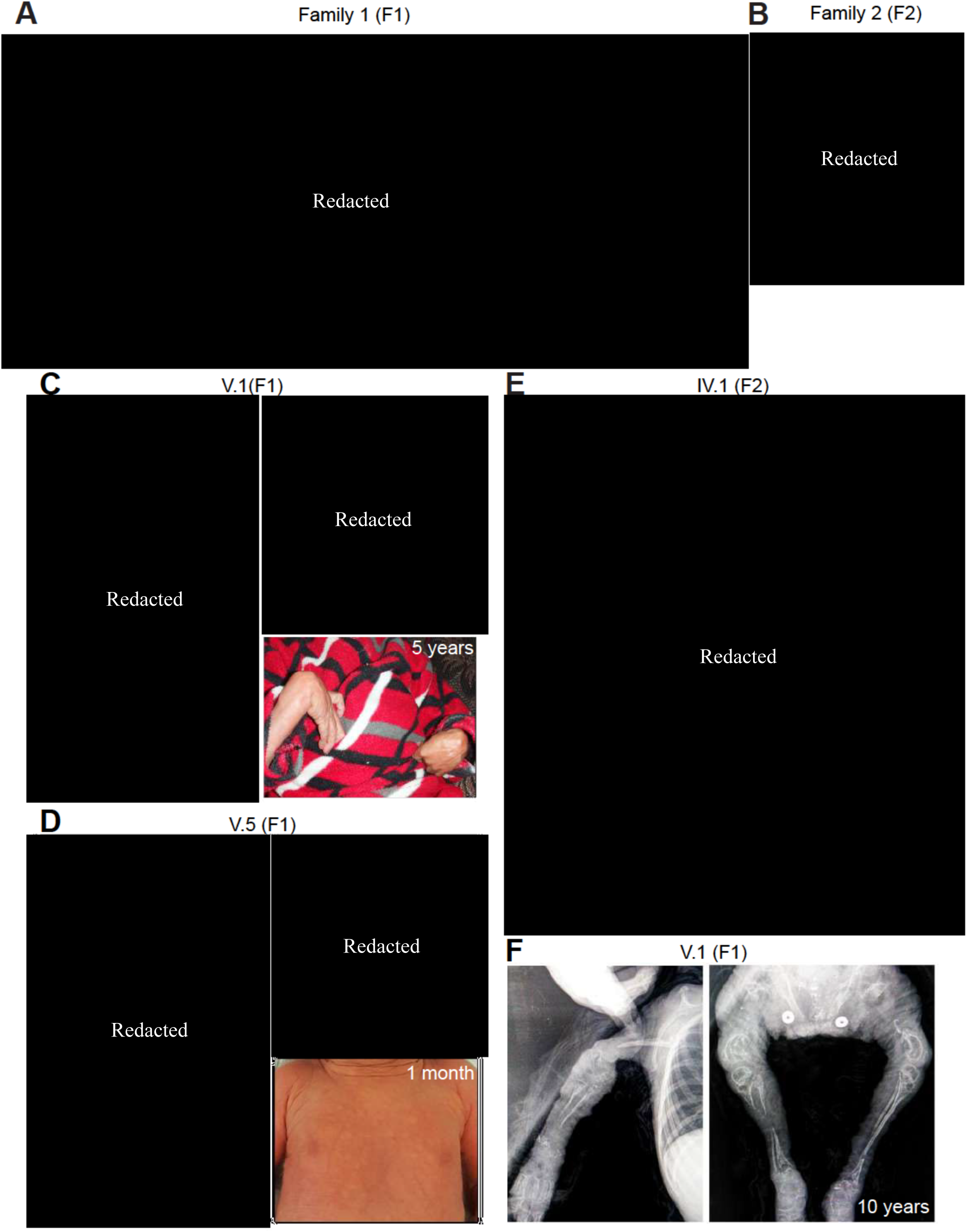
Patients from two distantly related families present with a recessively inherited syndrome characterized by osteogenesis imperfecta and neonatal progeria. (A, B) Pedigrees of two distantly related consanguineous families from 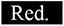, showing an autosomal recessive mode of inheritance of the disease. Black symbols and crossed symbols represent affected and deceased individuals respectively. (C-E) Pictures of investigated patients showing severe bone deformities and fractures, neonatal progeria, wrinkled skin, prominent forehead and pectus excavatum. (F) Radiographs of affected V.1 (F1) showing several deficits in the bones including deformity, dysplasia, spared joints and evidence of previous fractures. Severe calcification defects can also be noticed, involving premature atherosclerotic vascular calcification, periarticular soft tissue calcification and irregular calcification of carpal bones. The pedigrees and patients’ images have been removed per medRxiv policy on identifying information.

### A deep intronic *TAPT1* variant segregates with the disease

Although the two families were reported to be unrelated, both originated from 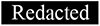 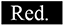. Assuming a founder mutation, we carried out homozygosity mapping for a total of 15 individuals including 3 affected patients (V.1 and V.5 (F1), IV.1 (F2)), 3 pairs of parents, and 6 unaffected siblings from F1 (IV.4, IV.5, IV.6, V.2, V.3 and V.4). Homozygosity mapping confirmed distant relatedness between the 2 families with a minimal shared locus on chromosome 4 (4p16.1-p15.31) (hg19). The length of this Identical-by-Descent (IBD) locus was 8.4 Mb spanning a total of 39 candidate genes (Fig. 2A, S2A). We first performed whole-exome sequencing (WES) for V.1 (F1) and IV.1 (F2), but no compelling recessive mutations were found. To expand our search, we turned to an unbiased RNA-seq approach using primary cutaneous fibroblasts from 2 affected individuals (V.1 and V.5 (F1)), and 2 unrelated wildtype (WT1 and WT2) controls. Out of the 39 candidate genes in the IBD region, our differential expression analysis data disclosed that *TAPT1* was the only significantly dysregulated transcript in the patient primary dermal fibroblasts (Log2 fold change = -2.5) (Fig. 2B, S2B, S2D). Moreover, the alternative splicing analysis identified 63 genes with splicing defects in patient samples, *TAPT1* being the top significant transcript with an exon 12 skipping event (Fig. 2C, 2D, S2C). Interestingly, homozygous mutations in *TAPT1* were previously reported as the genetic cause of complex osteochondrodysplasia (MIM616897)^14^. Although less severe, this disease bears strong clinical overlap with the syndrome reported in this study. The exon 12-skipping event prompted us to search for the presence of possible *TAPT1* intronic mutations. Targeted Sanger sequencing for this exon and its neighboring nucleotides (∼300 bps) disclosed a deep intronic single nucleotide polymorphism (c.1237-52 G>A) within intron 11 that entirely segregated with the disease in all available family members (Fig. 2D, 3A). Together, these findings indicate that the c.1237-52 G>A mutation within intron 11 of *TAPT1* most likely caused the disease for the 6 affected children.

**Figure 2.**
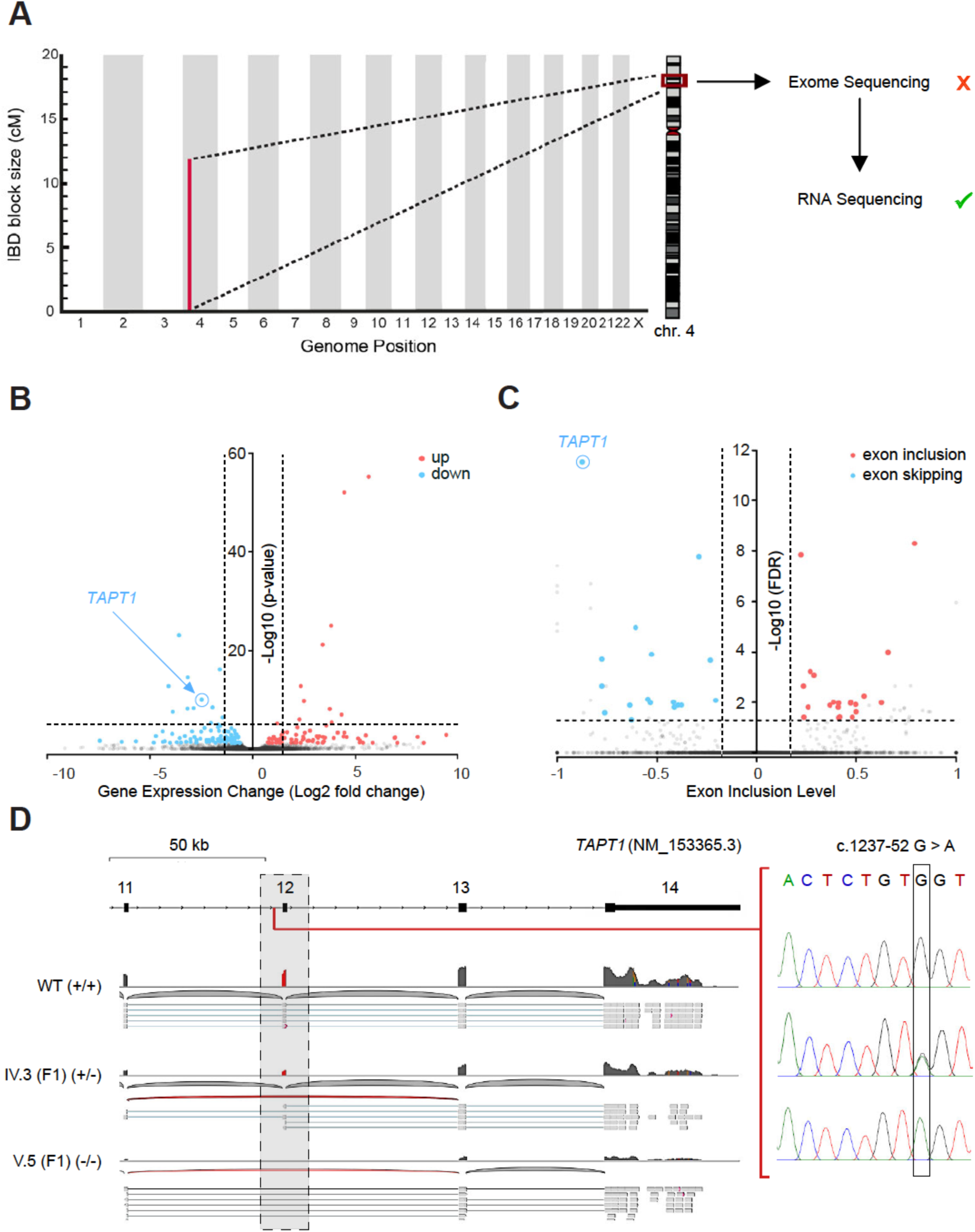
Homozygosity mapping followed by RNA-seq uncovers a deep intronic recessive mutation in *TAPT1*. (A) Schematic representation of the shared IBD region between both 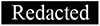 families, located on Chromosome 4 (4p16.1 - p15.31) with a size of ∼12 cM. Whereas WES analysis did not reveal any mutations in the coding sequences located in the IBD region, RNA-seq analysis helped us to identify the disease causative gene from this locus. (B) Volcano plot showing differentially expressed genes between WT (WT1 and WT2) and patient (V.1 (F1), V.5 (F1)) primary dermal fibroblasts. The vertical axis (*y*-axis) shows the –log10 P-value, whereas the horizontal axis (*x*-axis) displays the log2 fold change value. The red dots represent the upregulated transcripts; the blue dots represent the downregulated transcripts. A total of 172 genes were found significantly dysregulated. *TAPT1*, a gene located in the IBD region, appeared among the most significantly downregulated genes in the patients. (C) Plot showing the alternative splicing analysis results from WT (WT1 and WT2) and patient (V.1 (F1), V.5 (F1)) primary dermal fibroblasts. The vertical axis (*y*-axis) shows the –log10 FDR (False Discovery Rate), whereas the horizontal axis (*x*-axis) represents the exon inclusion level (value ranging from -1 to 1). The red dots represent transcripts with exon inclusion events; the blue dots represent transcripts affected by exon skipping. A total of 63 aberrantly spliced genes were found in the patient cells, being *TAPT1* the most significant exon skipping event. (D) (Left) Schematic representation showing the complete loss of exon 12 from *TAPT1* transcript in patient cells, as defined by our splicing analysis data. (Right) Chromatogram showing the novel intronic mutation (c.1237-52 G>A) we found entirely segregating with the disease in all available family members. For display purposes, results from the targeted Sanger sequencing in WT, IV.3 (F1) and V.5 (F1) individuals are shown. The mutation is present in heterozygosis in IV.3 (F1) (unaffected mother) and in homozygosis in V.5 (F1) (affected patient).

**Figure 3.**
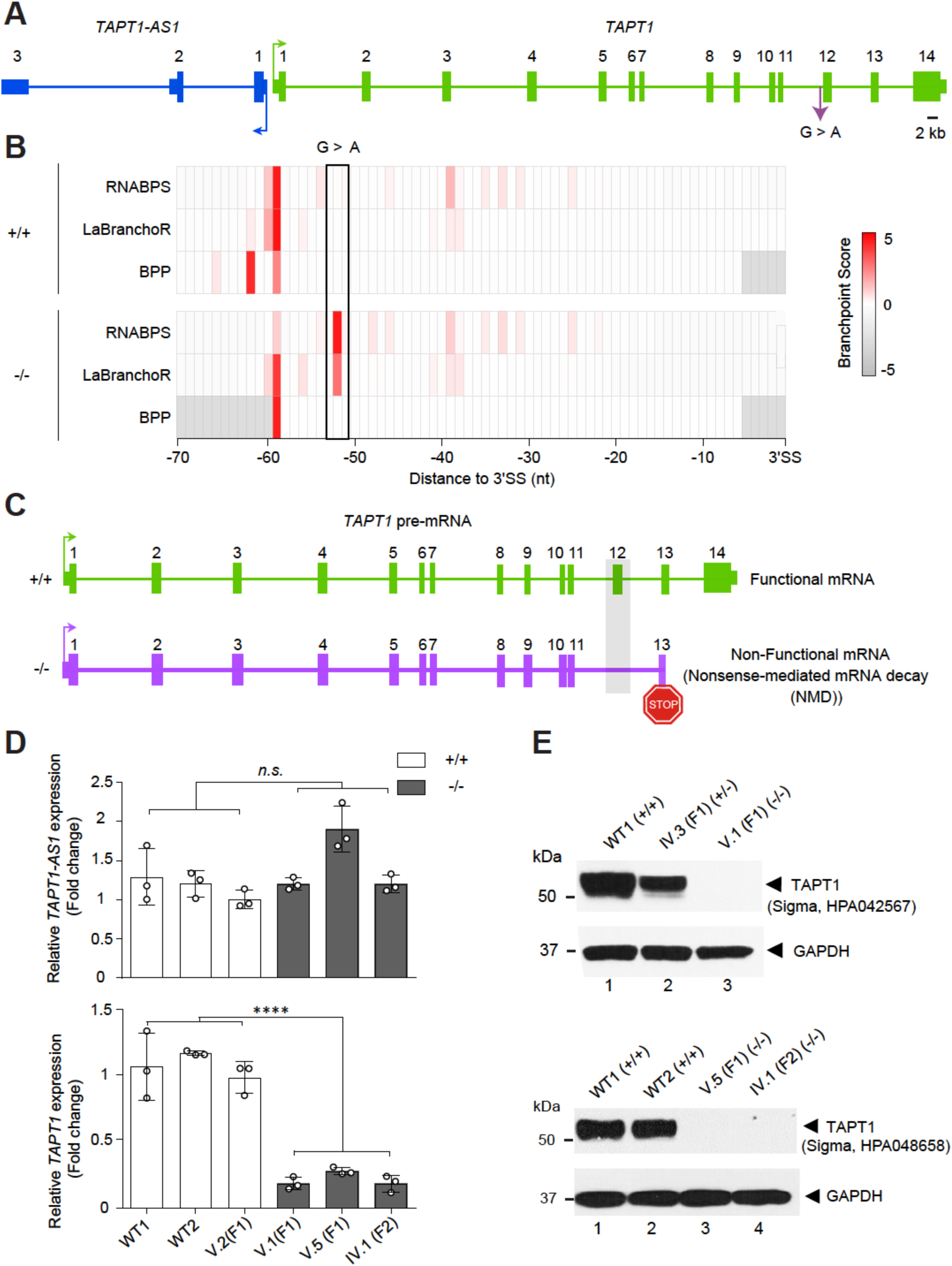
*TAPT1* mutant transcript lacks exon 12 and undergoes NMD to create a protein-null allele. (A) Schematic representation of *TAPT1* and *TAPT1-AS1,* indicating the causative intronic mutation (c.1237-52 G>A). The transcription start sites and the direction of transcription are indicated by arrows. Scale bar represents 2 kb. (B) Diagram showing the branchpoint scores for the target c.1237-52 position and flanking nucleotides in *TAPT1* intron 11 in both WT (+/+) and patient cells (-/-), as obtained from the RNABPS ^69^, LaBranchoR ^70^ and BPP ^71^ softwares. High branchpoint scores were predicted for the G>A transition in the patient cells using the RNABPS and LaBranchoR methods. The *x*-axis represents the nucleotide distance to the 3’ splice site (3’SS). (C) Schematic representation showing that the complete loss of exon 12 in *TAPT1* results in a premature stop codon, which targets the transcript for nonsense-mediated mRNA decay. (D) qPCR results using specific primers for *TAPT1* and *TAPT1-AS1* in 3 WT (WT1, WT2 and V.2 (F1)) and 3 affected (V.1 (F1), V.5 (F1), IV.1 (F2)) individuals. *TAPT1* mRNA is significantly reduced in all patients compared to WTs, whereas *TAPT1-AS1* transcript levels are unaffected. Fold change relative to V.2 (F1) is plotted as mean ± SD. Asterisks indicate conventional statistical significance (Student t-test; n.s. p-value > 0.05, **** p-value < 0.0001). (E) Western blot analysis of endogenous TAPT1 protein (∼60 kDa) using whole protein extracts from primary dermal fibroblasts from WT (WT1 and WT2), heterozygous (IV.3 (F1)) and homozygous (V.1 (F1), V.5 (F1) and IV.1 (F2)) individuals and two different commercial antibodies (top: Sigma, HPA042567; bottom: Sigma, HPA048658). Results show a complete absence of TAPT1 protein in patient samples. GAPDH was used as a loading control.

### Exon 12 skipping targets *TAPT1* mutant transcript for NMD, resulting in a protein-null allele

How does the deep intronic mutation in *TAPT1* lead to disease? To gain insights into the underlying disease-causing molecular mechanism, we applied a combined computational and experimental approach. We found that the private homozygous c.1237-52 G>A transition was predicted to serve as an alternative splicing branchpoint (Fig. 3B), thereby resulting in the exclusion of *TAPT1* exon 12 (Fig. 2D, 3C). As the complete loss of exon 12 creates a premature stop codon, we used orthogonal qPCR validation tests to investigate whether the mutant *TAPT1* transcript is targeted for nonsense-mediated decay (NMD) (Fig. 3C). Our data confirmed the statistically significant reduction of endogenous *TAPT1* mRNA levels in three of the patients compared to WT individuals (Fig 3D). We next examined the effect of the identified deep intronic *TAPT1* mutation on the expression of its encoded protein. We employed two different commercial antibodies to detect endogenous TAPT1 in protein extracts from primary fibroblast cultures from two distinct patients and two WT individuals. Western blotting with both antibodies showed a complete loss of endogenous TAPT1 protein in all patient cells carrying the c.1237-52 G>A mutation in homozygosity (Fig. 3E). Notably, the fibroblasts from the mother IV.3 (F1) showed intermediate TAPT1 protein levels, which were consistent with her heterozygous genotype (Fig. 3E). These findings indicate that the novel mutant variant reported in this study behaves as a protein-null allele by creating an aberrant mis-spliced *TAPT1* transcript, which undergoes degradation before being translated.

*TAPT1* is situated head-to-head with its sequence-related antisense gene *TAPT1-AS1* (Fig. 3A), which encodes a long non-coding RNA. Such upstream antisense transcripts can play a critical role in the regulation of gene expression ^16–19^, in particular towards their associated protein-coding genes ^20, 21^. Here, because of the manifest physical proximity of *TAPT1* and *TAPT1-AS1*, it is likely that both genes are expressed in a coordinated manner through shared regulatory elements as previously described for the majority of long non-coding RNA:mRNA gene pairs ^22^. As such we examined whether the downregulation of *TAPT1* may also alter the expression levels of *TAPT1-AS1.* qPCR data showed no overt changes in *TAPT1-AS1* levels in patients’ fibroblasts relative to control cells (Fig. 3D), thereby indicating that TAPT1 downregulation does not affect the expression of its neighbor antisense transcript. To further investigate the possible regulatory function of *TAPT1-AS1* on its target gene, we knocked down the endogenous transcript in WT and TAPT1 mutant fibroblasts by transient transfection of two different *TAPT1-AS1* GapmeRs. As evidenced in our qPCR results, although both GapmeRs achieved the near complete depletion of *TAPT1-AS1* transcript in both control and patient cells, no significant alterations were detected in *TAPT1* mRNA levels (Fig. S3A). In addition, TAPT1 protein expression was also found unaffected in a *TAPT1-AS1* knocked down context (Fig. S3B). These results argue against the potential regulatory role of *TAPT1-AS1* on *TAPT1* expression, and hence exclude the possibility that this antisense transcript could be having an impact on the pathogenesis of the disease found in our patients.

### TAPT1 is enriched in the ER/Golgi and is dispensable for HCMV gH infection

*TAPT1* (MIM#612758) codes for a protein termed Transmembrane Anterior Posterior Transformation 1, with 5 membrane-spanning helices (Fig. 4A). Its cellular localization has been reported to be either in the endoplasmic reticulum (ER) or at the centrosome ^11, 12, 15^. In order to gain further insights in its cellular localization, we performed immunofluorescence (IF) staining with both commercial TAPT1 antibodies in WT and mutant primary dermal fibroblasts. Each antibody yielded different staining patterns which were identical between control and TAPT1 knockout cells (Fig. S4A, S4B). This clearly indicated that these antibodies are not suitable for IF purposes. Since we validated the use of the same antibodies for western blotting, we opted to conduct subcellular fractionation on patient and WT fibroblasts as an alternative strategy to examine its subcellular localization. Endogenous TAPT1 was enriched in the Mito/ER/Golgi fractions and, to a lesser extent, in the nuclear fractions (Fig. 4B). This data is consistent with the previous report of EMP65, the homologous TAPT1 protein in yeast ^11, 12^, and pfcarl, the homologous TAPT1 protein in plasmodium ^9^, localizing in the ER/Golgi apparatus. Additional evidence of localization of TAPT1, with its partner protein SUCO, in the secretory pathway was also obtained in various human cell lines ^23^.

**Figure 4.**
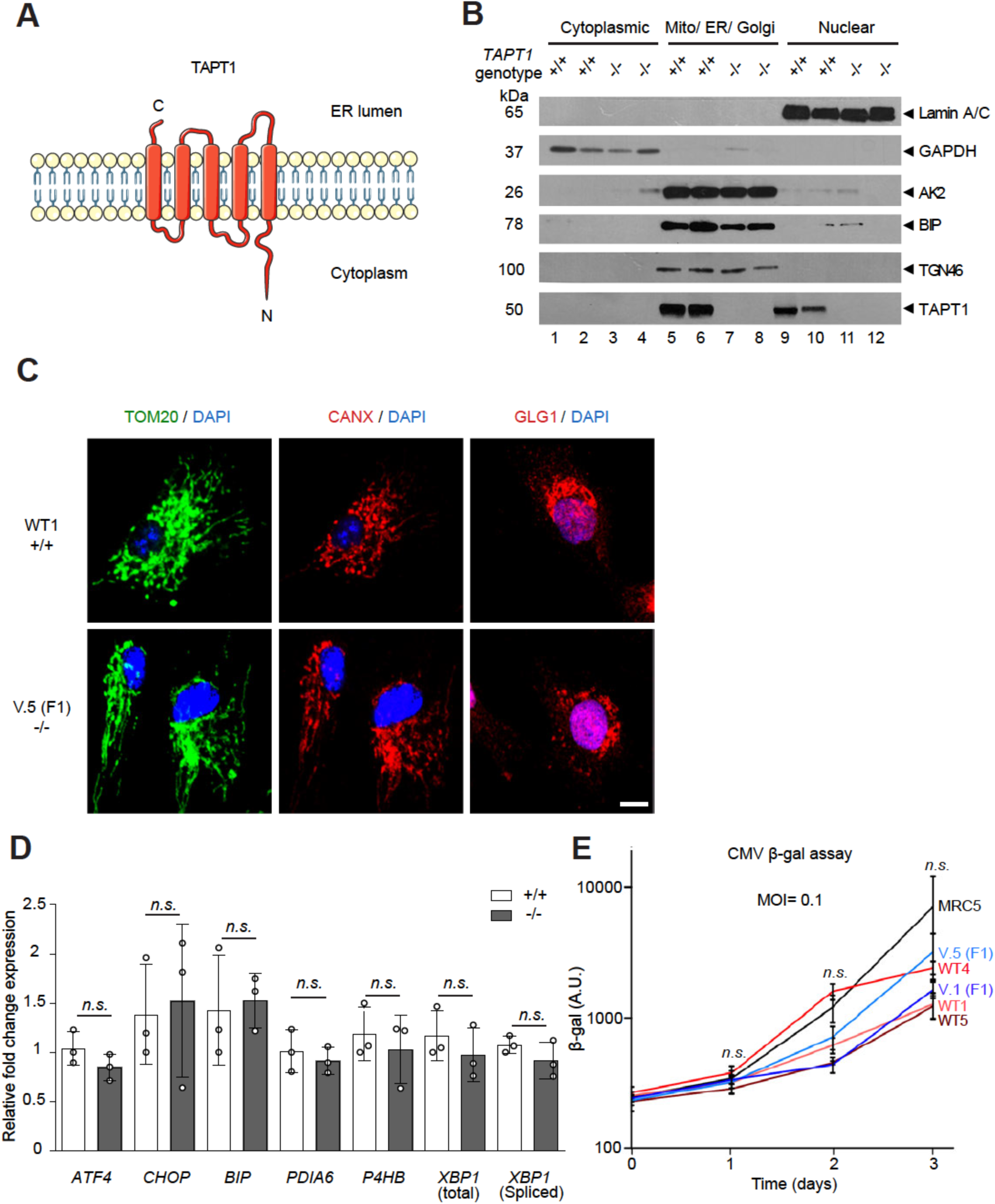
TAPT1 cellular localization and functional data. (A) TAPT1 predicted topology: a membrane-spanning protein consisting of 5 transmembrane helices (Uniprot database). (B) Western blot analyses for TAPT1 (∼60 kDa) using cytosolic, Mito/ER/Golgi and nuclear protein extracts from primary dermal fibroblasts of two WTs (WT1 and WT2) and two patients (V.5 (F1) and IV.1 (F2)). TAPT1 protein is highly enriched in the Mito/ER/Golgi fraction, and to a lower extent in the nuclear fraction. GAPDH, TGN46 and BiP served as a cytosolic, Golgi network and ER markers, respectively. Adenylate Kinase (AK2) was used as a mitochondrial marker. Laminin A/C was used as a nuclear marker. (C) Immunofluorescence staining of mitochondria using anti-TOM20 (green), ER using anti-CANX (red) and Golgi using anti-GLG1 (red) in primary dermal fibroblasts from WT1 and V.5 (F1). Similar staining patterns are observed with the three antibodies in both cell lines. Scale bar represents 10 µm. (D) qPCR analysis of a panel of canonical ER stress markers shows no significant differences in 3 patients (-/-) (V.I (F1), V.5 (F1) and IV.1 (F2)) primary dermal fibroblasts compared to WTs (+/+) (WT1, WT2 and WT3) cells. Fold change relative to WT is plotted as mean ± SD. Statistical significance was tested by Student t-test (n.s. p-value > 0.05). (E) CMV cell infection assay on 2 patient (V.1 (F1) and V.5 (F1)) and 3 WT (WT1, WT4 and WT5) primary dermal fibroblast cell lines, using β-galactosidase activity as a readout. MRC5 cell line was used as a positive control. All of the cells were infected by the HCMV strain RC256 at a MOI=0.1. Data are shown as mean ± SD. Statistical significance was tested by Student t-test (n.s. p-value > 0.05).

We carried out a series of functional tests by comparing WT and patient fibroblasts in order to gain a better understanding of TAPT1’s cellular function. Several studies in yeast have reported that EMP65 is critically involved in the Unfolded Protein Response (UPR) ^11^ and ER-Associated Degradation (ERAD) pathways ^12^. However, we could not document significant alterations in the expression levels of a panel of ER stress-associated markers in *TAPT1* knockout cells by qPCR (Fig. 4D). IF staining did not reveal obvious defects in ER-(using an anti-CANX antibody), GOLGI- (using an anti-GLG1 antibody) or mitochondrial-morphology (using anti-TOM20 antibody) in patient TAPT1 knockout cells compared to WT cells (Fig. 4C).

Two early publications had reported that *TAPT1* encodes for a receptor of the human cytomegalovirus (HCMV) gH ^13, 14^. We revisited this claim by testing whether human TAPT1-null patient cells were resistant to HCMV strain RC256 infection. HCMV strain RC256 is a recombinant virus carrying the *Escherichia coli lac*Z gene as a marker under the control of the β gene promoter ^24^. Our β-gal reporter assay showed no discernable differences between WT and mutant patient cells (Fig. 4E), hence indicating that TAPT1 is not essential for HCMV infection and suggesting the likely presence of other cellular receptors which permit HCMV cellular entry in the absence of TAPT1.

### Extracellular matrix and collagen-related pathways are dysregulared in TAPT1-null cells

To gain insights into the cellular role of TAPT1 and the disease mechanism, we investigated which genes and pathways were changed in TAPT1-null patient cells. The combined analysis of RNA-seq and SI-NET-seq data was highly informative in this context. RNA-seq quantifies the steady-state RNA levels whereas SI-NET-seq provides a quantitative measure of the Pol II occupancy with single-nucleotide precision genome-wide ^25^. SI-NET-seq is an improved variant of the NET-seq approach ^26^ that relies on spike-ins and therefore allows quantitative comparisons between conditions ^25^. While the RNA-seq data identified *TAPT1* as the only dysregulated gene on the candidate Chr. 4 locus, it also provided an unbiased list of 172 significantly (p<0.05) dysregulated genes in the patients’ fibroblasts at the mRNA level (Fig. 5A) (Table S1). Of these significantly altered genes, a similar fraction was up- and downregulated (Fig. 5A) (Table S1). Beyond *TAPT1* which is the fifth most significantly downregulated gene in mutant cells, dysregulation of several other target genes was also validated by qPCR, such as *RARRES2*, *ZIC1*, and *ZIC4* (Fig. 5B). Moreover, SI-NET-seq results revealed a total of 317 genes with aberrant Pol II occupancy and dysregulated nascent RNAs in the patient cells (Fig. 5C, S5A, S5B) (Table S2). For the majority of these genes (70%) the density of transcriptionally engaged Pol II was significantly increased in patient cells (Fig. 5C) (Table S2). The Pol II occupancy at *TAPT1* was not changed in patient cells indicating that Pol II transcription of *TAPT1* was not impacted by the mutation (Fig. 5C). Importantly, the integrated analysis of RNA-seq and SI-NET-seq data provided a comprehensive view on the molecular pathways that were affected by the *TAPT1* mutation. A pathway analysis of genes with either a significantly altered mRNA level or Pol II occupancy consistently revealed that extracellular matrix (ECM) organization and collagen-related pathways were highly enriched (Fig. 5D), suggesting a role of TAPT1 in ECM and collagen dynamics.

**Figure 5.**
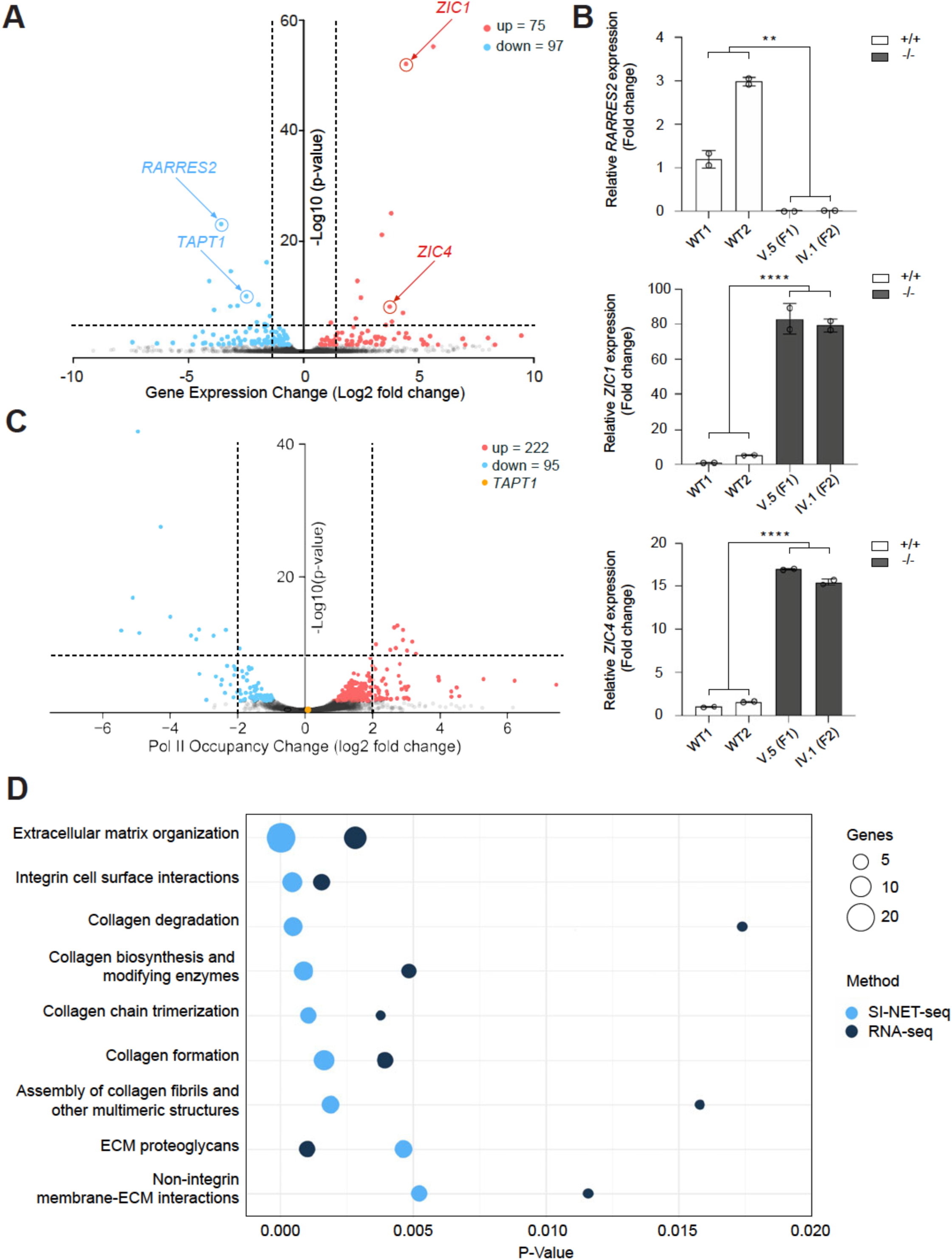
Integrated analysis of SI-NET-seq and RNA-seq data revealed enrichment of collagen and ECM-related pathways in TAPT1-null cells. (A) Volcano plot showing differentially expressed genes as determined by RNA-seq in patient primary dermal fibroblasts (V.1 (F1), V.5 (F1)) compared to WT (WT1 and WT2) cells. The y-axis shows the –log10 P-value, whereas the x-axis displays the log2 fold change value. The red dots represent 75 significantly upregulated genes, and the blue dots represent 97 significantly downregulated genes. (B) qPCR validation test for 3 top dysregulated genes (*RARRES2*, *ZIC1*, and *ZIC4*) detected by RNA-seq. The analysis was performed on primary dermal fibroblasts from 2 WTs (WT1 and WT2) and 2 patients (V.5 (F1) and IV.1 (F2)). Fold change relative to WT1 is plotted as mean ± SD. Asterisks indicate statistical significance (Student t-test; ** p-value < 0.01, **** p-value < 0.0001). (C) Volcano plot showing genes with an altered occupancy of transcriptionally engaged Pol II in patient (V.1 (F1), V.5 (F1)) compared to WT (WT1 and WT2) primary fibroblast cells. The y-axis shows the –log10 P-value, whereas the x-axis indicates the log2 fold change value for the Pol II occupancy. The Pol II density is increased in 222 genes (red dots), and decreased in 95 genes (blue dots). The yellow dot represents *TAPT1*. (D) Bubble plot showing enrichment of collagen and extracellular matrix (ECM) pathways from the integrated reactome pathway analysis ^72^ of the SI-NET-seq (light blue circles) and RNA-seq (dark blue circles) data. Enriched pathways are indicated on the y-axis, and the corresponding p-values are shown on the x-axis. The size of the circles represents the number of altered genes from each pathway.

## DISCUSSION

### Phenotypic spectrum of TAPT1 insufficiency

Here, we report the successful identification of a genetic variant causing a recessive Mendelian syndrome in six affected children presenting with severe bone defects, developmental delay and premature aging. As we did not detect any recessive mutations by WES, we followed an alternative analytical pipeline which involved homozygosity mapping, RNA-sequencing and targeted Sanger sequencing. We eventually identified a deep intronic mutation (c.1237-52 G>A) in the *TAPT1* gene that entirely segregated with the disease. It is known that pathogenic deep intronic mutations can induce splicing abnormalities, which in most cases lead to mRNA NMD due to the introduction of premature termination codons (PTCs) ^27–30^. Our prediction analysis suggested that the novel c.1237-52 G>A mutation is likely behaving as an alternative splicing branchpoint which triggers aberrant exon 12 skipping in the *TAPT1* pre-mRNA. This splicing aberration disrupts the reading frame and introduces a premature stop codon which targets mutant *TAPT1* transcript for NMD, as confirmed by qPCR in two distinct patient’s fibroblast cell lines. As expected and evidenced by western blotting data, the significant drop in *TAPT1* mRNA levels prevents the translation of a truncated protein. Moreover, SI-NET-seq showed that nascent transcription at the *TAPT1* gene was not affected by the mutation, confirming that the disease arises from a post-transcriptional dysregulation.

Genetic defects in *TAPT1* were firstly reported by S. Symoens *et al*. (2016) in two consanguineous families with a complex and lethal osteochondroplasia syndrome (MIM616897) ^15^. Furthermore, N. Patel *et al*. (2017) reported a homozygous truncating mutation (c.846+2insT) in *TAPT1* segregating with pediatric cataract, although these patients did not show any evidence of skeletal defects ^31^. In addition to the shared clinical features with these previously reported TAPT1-deficient patients including bone abnormalities and cataract, our affected children also suffered from neonatal progeria, characterized by wrinkled and thin skin, premature depigmentation and lipodystrophy. This vast phenotypic variation may be driven by the severity of the alleles identified. Our six new patients carry a complete loss-of-function mutation resulting in a protein-null allele, whereas both prior studies showed partial loss-of-function mutations including missense and in-frame exon 6 and exon 10 skipping ^15, 31^. Another possibility is that, as is the case for *LMNA* ^32^, a wide range of *TAPT1* diseases exist depending on which domain of the protein is mutated, thus accounting for the observed phenotypic heterogeneity.

### TAPT1-deficiency resembles a collagenopathy

To date, RNA-seq stands out as the gold-standard technique to identify affected signaling pathways underlying a certain disease. To identify cellular processes that are affected upon TAPT1 mutation, we performed an integrated pathway enrichment analysis combining RNA-seq and SI-NET-seq results. RNA-seq and SI-NET-seq uncovered genes with a significant change in transcript levels and nascent transcription in patient cells, respectively. Despite the different types of data, we observed a strong overlap in dysregulated pathways between both datasets. Collagen- and ECM-related pathways standout as most significant hits from this combined analysis indicating a dysregulation of these processes in patient cells. This interesting finding is consistent with our patients’ phenotype, which manifests with severe bone defects and skin abnormalities. Collagens are the most abundant proteins made by the human body and serve to provide structural support, tensile strength while mediating cell adhesion, and migration ^33, 34^. The bone tissue and the skin dermis account for 80% of the total collagen content of the body ^35^. Importantly, the majority of genetic alterations causing bone defects affect collagen themselves e.g. COL1A1 (MIM114000, MIM619115, MIM130060, MIM166200, MIM166210, MIM259420, MIM166220, MIM166710), COL1A2 (MIM619120, MIM617821, MIM225320, MIM166210, MIM259420, MIM166220, MIM166710), or enzymes dedicated to their processing and secretion such as P3H1 (MIM610915), CRTAP (MIM610682) and TANGO1 (MIM619269) ^36–38^. The clinical manifestations and transcriptomics results shown here support the hypothesis that TAPT1-deficiency belongs to the heterogeneous group of collagenopathies.

Previous computational and experimental interactome analyses proposed that TAPT1 physically interacts with two additional ER-resident proteins: SUCO (SUN Domain Containing Ossification Factor) ^23, 39^ and P4HB, also known as PDI1 (protein disulfide isomerase 1) ^39^. Notably, mutations in SUCO and P4HB have been linked to skeletal dysplasia ^40–45^ in humans, which aligns with the TAPT1 loss-of-function clinical presentation. *Tapt1* and *Suco* mutant mice successfully phenocopy their corresponding human disease as they also present with severe skeletal defects ^46, 47^. These two proteins form a highly conserved complex which is present in all eukaryotic cells from yeast ^11, 48^ to humans ^23^. EMP65 and SLP1, the yeast homologues for TAPT1 and SUCO respectively, have been shown to be involved in ER quality-control machinery including UPR ^11^ and ERAD pathways ^12^. Surprisingly, our functional analyses did not reveal major abnormalities in the ER morphology and expression levels of ER stress markers in *TAPT1*-null cells. Previous research actually reported unaltered protein levels of ER chaperones, including BIP/GRP78, Calnexin, and GRP94, in *Suco*-null mouse osteoblasts ^47^, which is consistent with our results assuming a common functional pathway for TAPT1 and SUCO. P4HB, the other proposed interacting partner for TAPT1, serves as a prototypic thiol isomerase that is involved in the hydroxylation of proline residues in collagen fibers ^49–51^. Therefore, our data adds to previous studies supporting the hypothesis of TAPT1, SUCO, and P4HB may form a functional complex residing in the ER/GOLGI and playing a key role in collagen post-translational processing with a particular relevance to skeletal development in vertebrate species. In accordance with this idea, delayed collagen folding and secretion was documented in *TAPT1* mutant fibroblasts ^15^.

### What could be TAPT1s’ universal function in eukaryotic cells?

Although loss-of-function mutations in *TAPT1, SUCO* and *P4HB* in humans all result in osteochondroplasia-like phenotypes, the homologues of these genes in lower organisms lead to unrelated phenotypes when absent. *POD1*, the TAPT1 homologue in *Arabidopsis*, was shown to be involved in pollen tube formation ^52^. *F26F2.7*, the *Caenorhabditis elegans* homologue of *TAPT1* is a critical gene for embryonic viability with undetermined function yet ^53^. In *Plasmodium falciparum,* the causative pathogen for malaria, mutations in *TAPT1’s* homologue, *pfcarl*, confer resistance to various structurally unrelated antimalarial compounds which appear to target the ER/Golgi function of the parasite ^10, 54, 55^. In the unicellular fungus *Saccharomyces cerevisiae*, TAPT1 which is known as Emp65, is required for the stability of soluble proteins that are targeted to the secretory pathway ^12^. Notably, none of these species of plants, invertebrates or fungus possess genes coding for collagens, arguing that TAPT1’s role in all eukaryotic cells must be unrelated to collagen biology *per se*, but instead fulfill a more essential cellular role.

## MATERIALS AND METHODS

### Sample Collection and Clinical Assessment

The affected children were firstly diagnosed 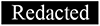 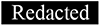 with severe osteogenesis imperfecta. In total, 15 saliva samples were collected from members of the two families including parents, affected and unaffected siblings. Genomic DNA from saliva samples was isolated using the Origene DNA Collection Kit (OG-500, DNAGenotek). Skin biopsies from three affected (V.I (F1), V.5 (F1) and IV.1 (F2)) and one unaffected (IV.3 (F1)) family members were also collected. Informed consent was obtained from all individuals in accordance with local ethical review board requirements in 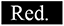 and Singapore (A*STAR IRB reference code #2019-087, Singapore). Additionally, parents gave their consent to participate in the present study and to have the results of this research work published.

### Genotyping and Homozygosity Mapping

SNP genotyping was performed on the genomic DNA from 15 affected and unaffected individuals from both families using Illumina HumanCoreExome-12v1 Bead-Chips. Identity-by-descent (IBD) mapping detected common homozygous regions in the 3 affected individuals using Wolfram Mathematica data-analysis software. IBD homozygous blocks were identified as regions >2 cM. Candidate homozygous regions were refined by excluding the shared homozygous regions with unaffected individuals. Finally, a single identical and homozygous region was revealed on Chr. 4 (4p16.1-p15.31) (hg19).

### Whole Exome Sequencing (WES)

The Ion TargetSeq^TM^ Exome and Custom Enrichment Kit (Life Technologies) was used for exome capture from 1 µg of genomic DNA from individuals V.I (F1) and IV.1 (F2). The Ion OneTouch System (Life Technologies) was used for exome library preparation. Sequencing was performed using the Ion Proton Instrument (Life Technologies) with one Ion PI chip (Life Technologies). The variants were annotated with their associated gene and location. No candidate variant was found using various filtering parameters.

### RNA-Sequencing

RNA from primary dermal fibroblasts from 2 patients (V.I (F1) and V.5 (F1)), and 2 unrelated wildtypes (WT1 and WT2) was extracted using the RNeasy Mini Kit (Qiagen). After measuring RNA quantity and integrity using the Agilent Bioanalyzer 2100 (Agilent Technologies), libraries were sequenced on a Illumina HiSeq/Novaseq sequencer. Reads were aligned to the GRCh38.p12 human reference genome using STAR v2.5.3a ^56^ with default parameters in paired-end mode.

For differential gene expression analysis, we quantified the transcript abundance of the annotated genes from GENCODE v28 ^57^ using HTSeq v0.11.4 ^58^ in ’union’ mode. Significant changes between the conditions were tested using DESeq2 v1.25.4 ^59^. We defined genes as significantly dysregulated when they had an FDR adjusted p-value of <0.05. For alternative splicing analysis, we focused on alternative exon inclusion and exclusion events between wildtype and patient samples. After read mapping, we identified all exons from GENCODE v28 annotation ^57^ showing an ‘exon inclusion level’ difference of at least 10% using rMATs v3.1.0 ^60^. The ‘exon inclusion level’ of an exon describes the fraction of reads accounting for the inclusion of the exon. We defined alternative exon usage as an event between conditions with a significant (FDR <0.05) difference in the ‘exon inclusion level’. Splicing events that were supported by less than 5 reads were excluded.

### SI-NET-sequencing

For spike-in NET-seq (SI-NET-seq) 15 x 10^6^ primary dermal fibroblasts were mixed with 3 x 10^6^ murine NIH 3T3 cells. Murine NIH 3T3 cells served as spike-in controls. All subsequent steps of the SI-NET-seq experiments were performed as recently described ^25^ with the following modification. For reverse transcription of nascent RNAs the SuperScript IV Reverse Transcriptase (ThermoFisher) was used.

Processing of SI-NET-seq data was performed as previously described ^25^ with some modifications. Briefly, adaptor sequences and unique molecular identifiers (UMIs) were trimmed by cutadapt v2.4 ^61^ and a custom python script, which preserves information of UMI sequences for the corresponding reads. The obtained reads were aligned to a joined reference genome from human GRCh38.p12 and mouse GRCm38.p6 using the STAR v2.5.3a aligner ^56^. For uniquely mapped reads, the position corresponding to the 3’-end of the nascent RNA fragment was recorded. We excluded reads that originated from reverse transcription mispriming and from PCR duplication using the UMI sequence information as described previously ^62^. Additionally, sequenced splicing intermediates were excluded. We masked regions that were transcribed by Pol I and III, as well as loci of short chromatin-associated RNAs, which were extracted from annotations in GENCODE v28/v29 ^57^ (mouse: M18 and M22), RefSeq v109 ^63^, miRBase v22.1 ^64^ and the UCSC’s RepeatMasker ^65^. In the final step of data processing, we split the spiked-in mouse observations from sample observations.

We statistically tested the significance of changes in the Pol II occupancy. First, we quantified the Pol II occupancy at actively transcribed genes using SI-NET-seq data. Active genes had a calculated TPM value of at least one using RSEM v1.3.1 ^66^ quantifications from wildtype RNA-seq data. Second, we tested for significant changes in the Pol II occupancy using DEseq2 v1.25.4 ^59^. For data normalization we calculated the ‘Relative Log Expression’ on Pol II occupancy measurements from spiked-in mouse cells. Quantification of Pol II occupancy in mouse was calculated as for sample observations. To define actively transcribed genes, we used RNA-seq data available for NIH3T3 mouse cells (ENCODE: ENCSR000CLW) ^67^. Changes in the Pol II occupancy at genes with an FDR adjusted p-value of 0.05 or smaller were considered significant.

### Segregation Analysis

The position coordinates and sequence of the candidate gene were obtained from the UCSC database. The region of the candidate mutation was amplified by PCR from genomic DNA from all 15 individuals using specific primers. Direct Sanger sequencing was performed on the PCR products using the BigDye Terminator Cycle Sequencing Kit (Applied Biosystems). Primer sequences are shown in Table S3.

### Cell Culture

Primary dermal fibroblasts of affected and unaffected individuals were derived from skin biopsies following standard procedures ^68^. All human cell lines were cultured at 5% CO_2_ and 37°C in high glucose DMEM (HyClone) supplemented with 10% fetal bovine serum (FBS) (HyClone), 1X penicillin/streptomycin (Thermo Fisher Scientific) and 2 mM L-glutamine (Biological Industries), and tested negative for mycoplasm using the MycoAlert^TM^ Mycoplasma Detection Kit (Lonza). Murine NIH 3T3 cells (ATCC: CRL-1658) were grown in DMEM containing 10% FBS (Bovine Calf Serum, iron-fortified, Sigma) and 5% penicillin-streptomycin.

### Quantitative PCR

Total RNA was isolated from primary dermal fibroblasts using the RNeasy Mini Kit (Qiagen). RNA (1 µg) was reverse transcribed using the Iscript^TM^ cDNA Synthesis Kit (Bio-Rad) according to the manufacturer’s instructions. Transcript levels were assessed using the Power SYBR^TM^ Green PCR Master Mix (Applied Biosystems) and specific primers (Table S3) on the ABI Prism 7900HT Fast qPCR System (Applied Biosystems). qPCR assays involved three biological replicates per condition and three technical replicates per sample (N = 3, n = 3). *GAPDH* was used as the housekeeping gene to normalize gene expression.

### Western Blot

Total cellular protein extracts from primary dermal fibroblasts were obtained using RIPA buffer supplemented with 1X Protease Inhibitor Cocktail (Roche). Nuclear, Mito/ER/Golgi, and Cytoplasmic fractions were prepared using the Cell Fractionation Kit Standard (Abcam, ab109719) following the manufacturer’s instructions. Protein concentrations were measured using the Pierce^TM^ BCA Protein Assay Kit (Thermo Fisher Scientific). Samples were reduced in Laemmli loading buffer containing dithiothreitol, and denatured at 95°C for 5 minutes. Equal amounts of protein were loaded on precast 10% Tris/Glycine/SDS polyacrylamide gradient gels (Bio-Rad), followed by transferring on PVDF membranes (Bio-Rad) using the Trans-Blot® Turbo^TM^ Transfer System (Bio-Rad). Membranes were blocked in 5% milk in TBST for 1 hour at room temperature, and subsequently probed with the following primary antibodies diluted in 5% milk in TBST overnight at 4°C: mouse anti-GAPDH (1:1000; Santa-Cruz, sc-47724), rabbit anti-TAPT1 (1:1000; Sigma, HPA042567, reacted with TAPT1 sequence covering exon 13-14), rabbit anti-TAPT1 (1:1000; Sigma, HPA048658, reacted with TAPT1 sequence covering exon 6-8), mouse anti-BIP (1:1000; BD Biosciences, 610978), rabbit anti-TGN-46 (1:1000; Abcam, ab50595), rabbit anti-AK2 (1:1000; Proteintech, 11014-1-AP) and mouse anti-Lamin A/C (1:1000; EMD Millipore, MAB3211). After washes in TBST, secondary anti-mouse/HRP or anti-rabbit/HRP antibodies were used at 1:4000 dilution in 5% milk in TBST for 1 hour at room temperature. The signal was revealed with the SuperSignal^TM^ West Chemiluminescent Substrate System (Thermo Fisher Scientific, #34080/34076/34096) and developed using CL-Xposure^TM^ Films (Thermo Fisher Scientific) in a Carestream Kodak developer.

### Immunofluorescence Analysis

Primary dermal fibroblasts were cultured on 8-well glass chamber slides (Millicell EZ SLIDES) and fixed for 15 minutes in 4% paraformaldehyde in PBS at room temperature. The cells were permeabilized with 0.3% Triton-X100 in PBS for 15 minutes, and blocked in 1% BSA in PBS for 1 hour at room temperature. Samples were then incubated with the following primary antibodies diluted in 1% BSA in PBS overnight at 4°C: rabbit anti-TAPT1 (1:1000; Sigma, HPA042567), rabbit anti-TAPT1 (1:1000; Sigma, HPA048658), rabbit anti-TOM20 (1:1000; Proteintech, 11802-1-AP), rabbit anti-Calnexin (1:2000; Abcam, ab22595) and mouse anti-GLG1 (1:500; Abcam, ab103439). For visualization, 1:500 secondary antibodies conjugated to Alexa Fluor 568 or Alexa Fluor 488 (Invitrogen, Molecular Probes) were incubated for 1 hour at room temperature in the dark. 1 μg/ml DAPI (Life Technologies) was used for DNA staining, and cells were mounted using ProLong™ Diamond Antifade Mountant (Invitrogen). Images were captured using a FV1000 Olympus inverted confocal microscope equipped with a Leica camera.

### GapmeR Transfection

Primary dermal fibroblasts were seeded on 6-well plates at a density of 100,000 cells per well. The following day, cells were transfected using the Lipofectamine RNAiMAX Reagent (Invitrogen) with two GapmeRs specific for *TAPT1-AS1* and a non-targeted GapmeR as a negative control at a 40 nM concentration. The GapmeRs were purchased from Qiagen (Germany), and their sequences are given in Table S3. 72 hours post-transfection, RNA and protein were harvested for downstream experiments.

### CMV β-Gal Assay

20,000-40,000 cells were plated per well in 96-well plates. MRC5 (human lung fibroblast cells) was also used as a positive control. The next day after seeding, the virus (CMV strain RC256 ATCC VR-2356) was added in DMEM supplemented with 10% FBS at MOI=0.1 and absorbed for 1 hour at room temperature. Then the virus was aspirated off and the plates were carefully washed twice with PBS. 80 µl of DMEM supplemented with 10% FBS were added back to each well and the plate was returned to the incubator. β-gal activity was read at different time points (Days 0, 1, 2, 3), involving three replicates per time point and per cell line. For that purpose, 20 µl of 5X lysis buffer (500 mM K-phos pH 7.8, 1% Triton X-100) were first added to each well. After pipetting up and down, samples were incubated for 15 minutes at 37°C. Then, 10 µl of the lysates were transferred to a plate with Galacto-Star, which was subsequently covered with foil and incubated at room temperature for 20 minutes. The signal was finally measured in a luminescence plate reader.

## Data Availability

All data produced in the present study are available upon reasonable request to the authors

## ACKNOWLEDGEMENTS

We are profoundly grateful to all patient family members for their participation in this study. We would like to thank all members of Reversade laboratory for their kind help and support. We would also like to thank Dr. Shokouh Karimi for her help in careful clinical evaluation.

## AUTHOR CONTRIBUTIONS

N.N. and B.R. designed the functional studies. M.E.K, R.F, W.S, A.A. and M.S. made clinical diagnoses and collected patient samples. C.B. performed homozygosity mapping and WES. A.B, A.M. and S.F. performed and analyzed RNA-seq and SI-NET-seq. S.D. and J.L.C performed HCMV experiments. N.N, R.M.T, P.H.C, N.E.B and Z.H. performed the functional and biomedical experiments in patient cells. N.N, A.M, A.B, P.H.C, R.M.T and B.R wrote the manuscript.

---

**Figure S1.**
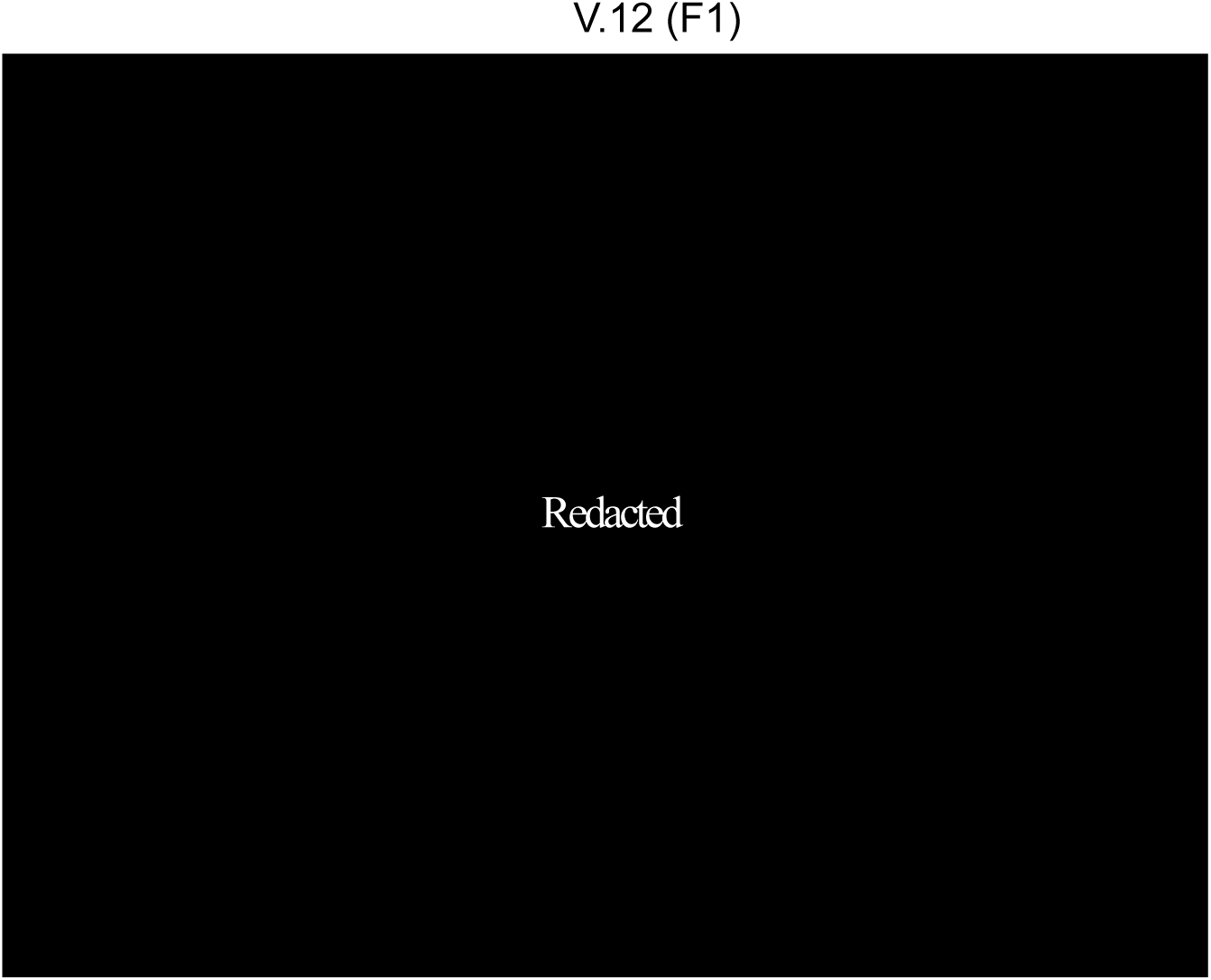
Clinical pictures of the affected V.12 (F1) individual. The patient presented with multiple abnormalities including bone and joint deformities, pectus excavatum, plagiocephaly microphthalmia and bilateral hypotropia. Moreover, she had apparent dysmorphic facial features such as a depressed nasal bridge and low set of ears. The patient’s images have been removed per medRxiv policy on identifying information.

**Figure S2.**
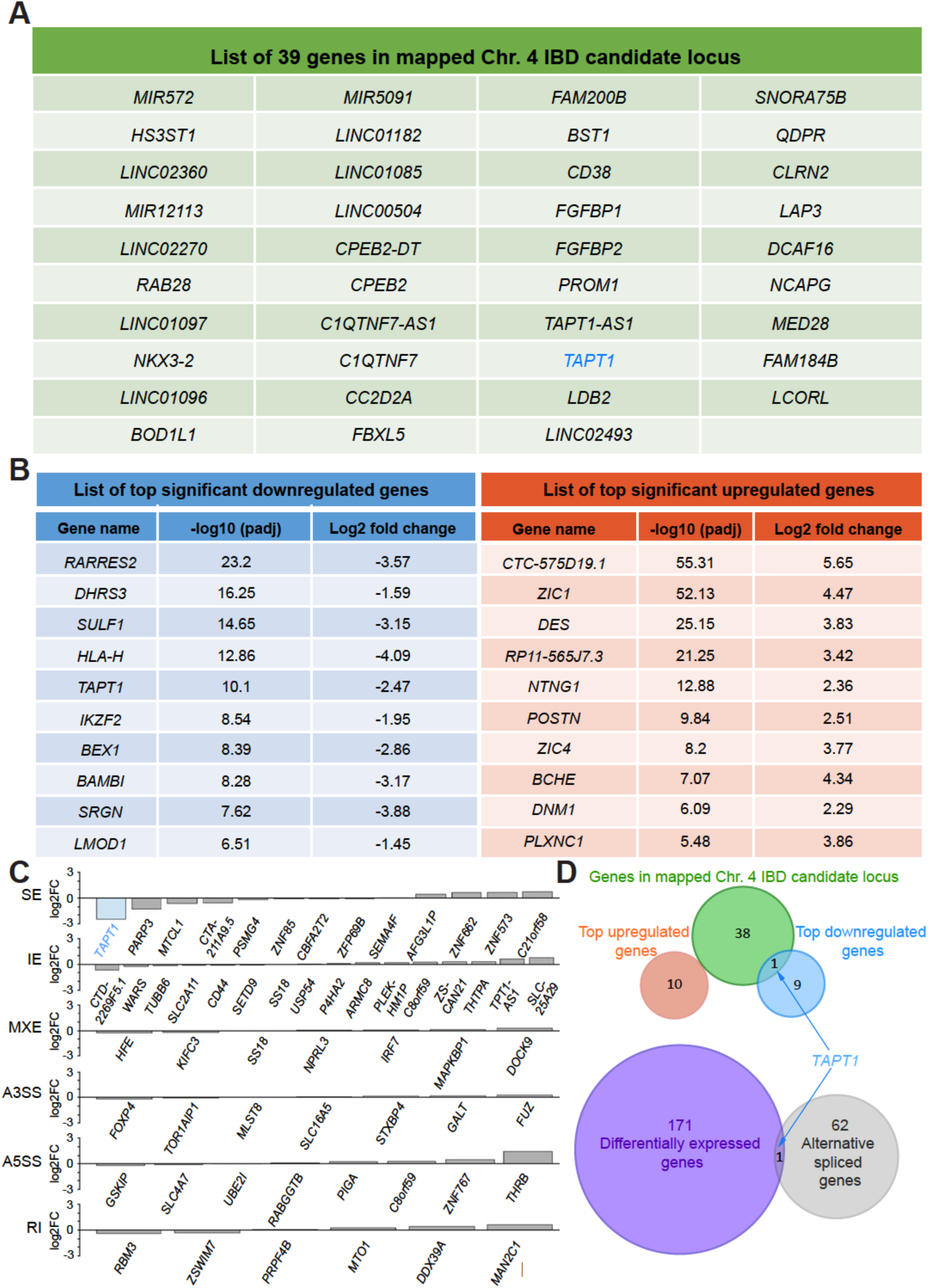
Overlap analysis from homozygosity mapping and RNA-seq data revealed TAPT1 as the only candidate gene. (A) List of the 39 candidate genes located in the mapped Chr. 4 IBD locus. (B) Lists of the top 10 significantly downregulated (left, blue) and upregulated (right, red) genes obtained from our RNA-seq differential expression analysis. (C) Expression changes (x-axis, log2FC) for genes with at least one alternative splicing event (skipped exon (SE), retained exon (RE), mutually exclusive exon (MXE), alternative 3’ or 5’ splice site (A3SS and A5SS) and retained intron (RI)). (D) (Top) Venn diagram displaying overlapping genes between the Chr. 4 IBD candidate locus, and the top 10 upregulated and downregulated genes from our RNA-seq data analysis. (Bottom) Venn diagram showing the overlapping genes between the differentially expressed set and the alternative spliced set from our RNA-seq data analysis. TAPT1 appears as the only overlapping gene in both diagrams.

**Figure S3.**
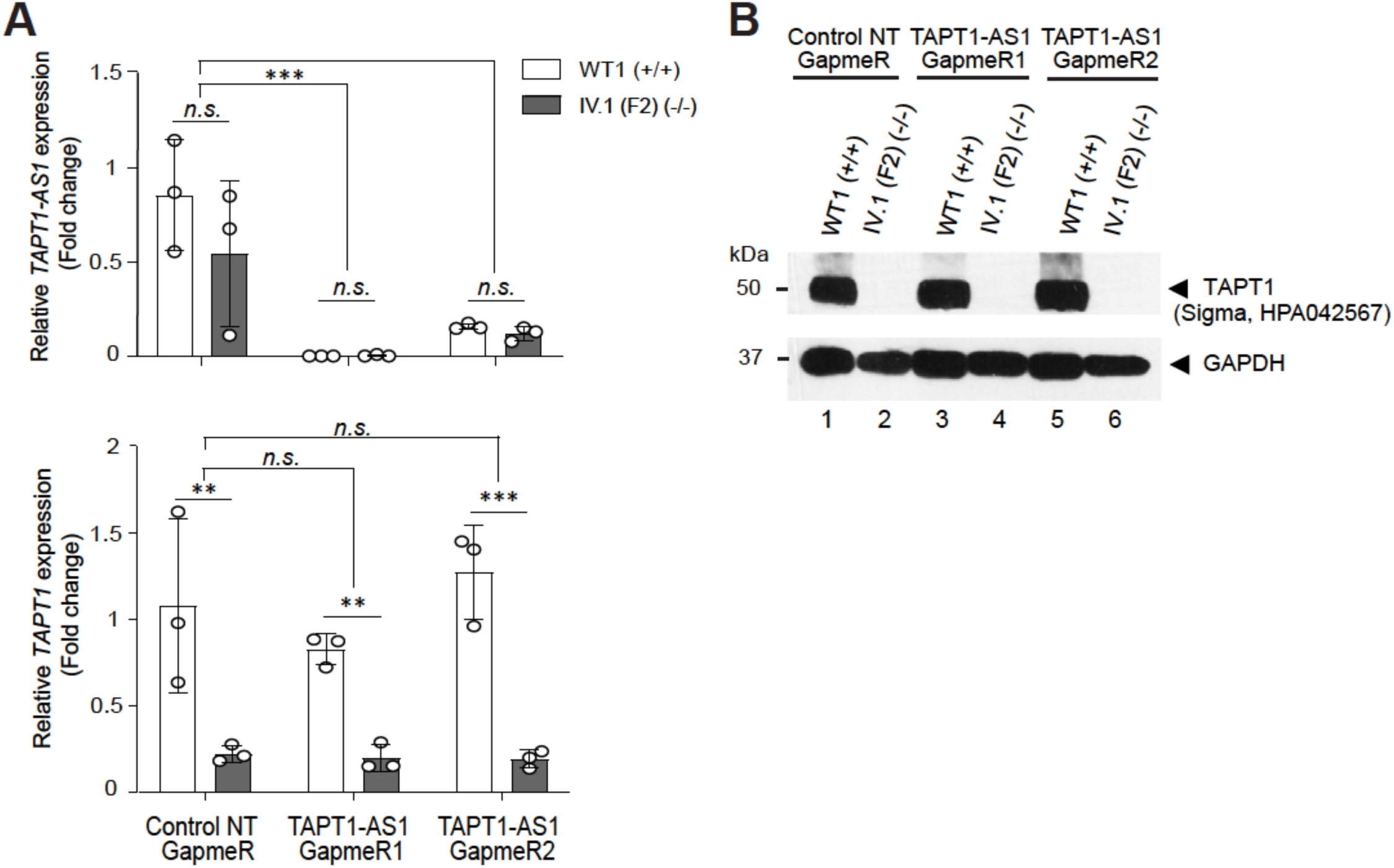
*TAPT1-AS1* shows no observable regulatory activity on *TAPT1* gene expression. Knockdown of *TAPT1-AS1* transcript using two different GapmeRs (1 and 2) in WT (WT1) and patient (IV.1 (F2)) primary dermal fibroblasts. A non-targeted (NT) GapmeR was used as control. (A) qPCR analysis of *TAPT1-AS1* (top) and *TAPT1* (bottom) transcript levels in the GapmeR-transfected cells. Results show the successful knockdown of *TAPT1-AS1* by both GapmeRs 1 and 2 compared to the control NT GapmeR. However, *TAPT1* mRNA levels are unaltered in both WT and patient cells. Fold change relative to WT1-Control NT GapmeR is plotted as mean ± SD. Asterisks indicate conventional statistical significance (Student t-test; n.s. p-value > 0.05, ** p-value < 0.01, *** p-value < 0.001). (B) Western blotting of protein extracts from the GapmeR-transfected cells, probing for TAPT1 (Sigma, HPA042567 antibody). Data shows that TAPT1 protein levels are unaffected by the knockdown of *TAPT1-AS1*. GAPDH was used as loading control.

**Figure S4.**
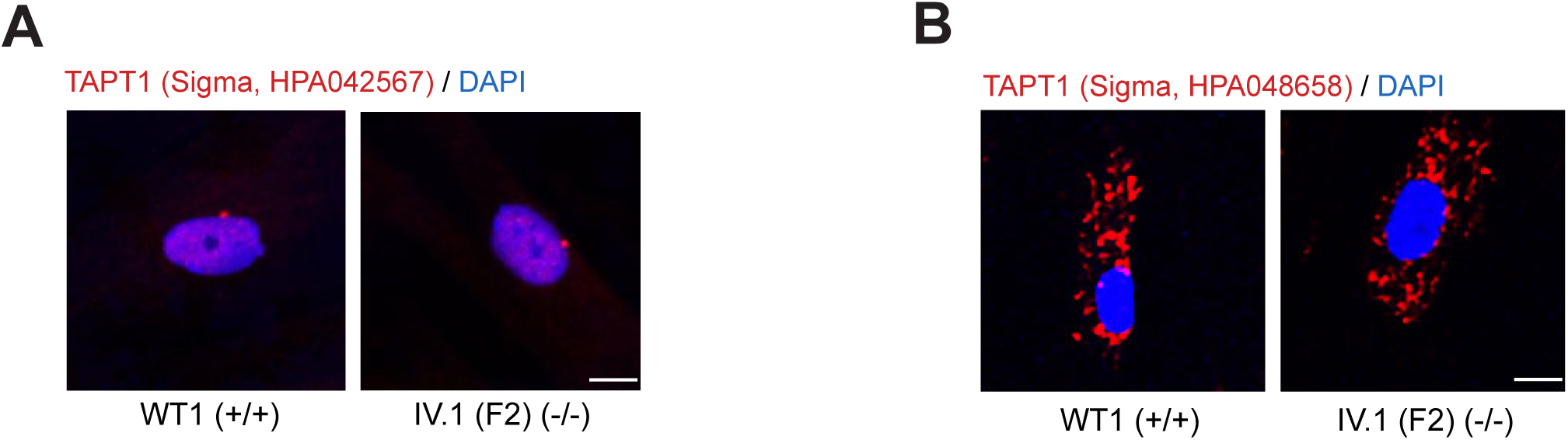
TAPT1 commercial antibodies are unsuitable for immunofluorescence experiments. Immunofluorescence staining using two different TAPT1 commercial antibodies (A: Sigma, HPA042567; B: Sigma, HPA048658) in WT1 and IV.1 (F2) primary dermal fibroblasts. Similar fluorescent signal was detected in WT and TAPT1-null cells in both cases. Scale bar represents 10 µm.

**Figure S5.**
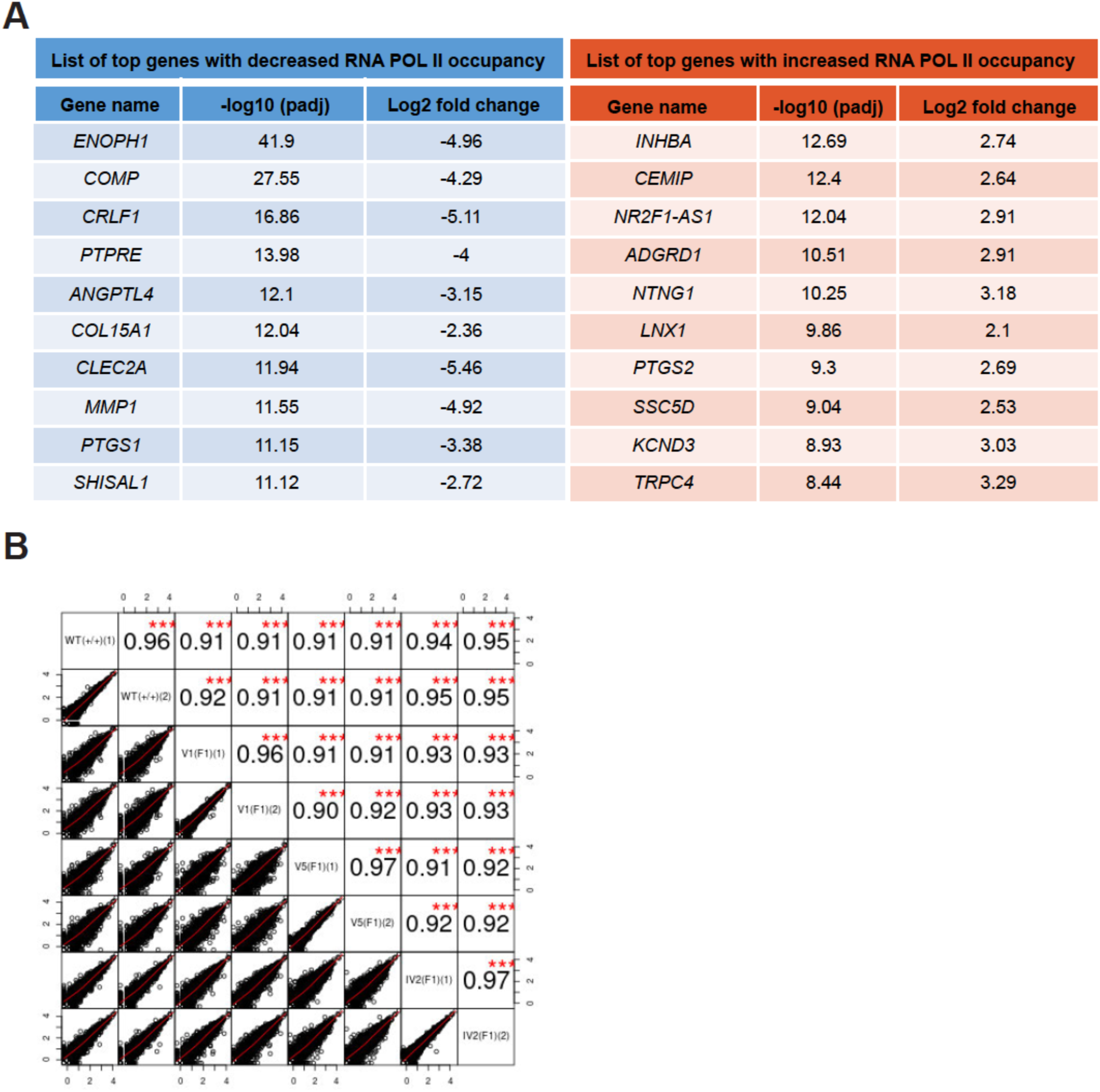
NET-seq analysis data. (A) Lists of the top 10 genes with significantly decreased (left, blue) or increased (right, red) RNA Pol II occupancy from our NET-seq analysis. (B) High Pearson’s correlation coefficients (r ≥ 0.96) between replicates of Pol II gene occupancy indicate the reproducibility of SI-NET-seq measurements.

## References

1. Jamshidi F, Place EM, Mehrotra S, et al. Contribution of noncoding pathogenic variants to RPGRIP1-mediated inherited retinal degeneration. Genet Med 2019; 21: 694–704.

2. Khan AO, Becirovic E, Betz C, et al. A deep intronic CLRN1 (USH3A) founder mutation generates an aberrant exon and underlies severe Usher syndrome on the Arabian Peninsula. DOI: 10.1038/s41598-017-01577-8.

3. Chen J, Ma N, Zhao X, et al. A rare deep intronic mutation of PKHD1 gene, c.8798-459 C > A, causes autosomal recessive polycystic kidney disease by pseudoexon activation. J Hum Genet 2019; 64: 207–214.

4. Djebali S, Davis CA, Merkel A, et al. Landscape of transcription in human cells. Nature 2012; 489: 101–108.

5. Chong JX, Buckingham KJ, Jhangiani SN, et al. The Genetic Basis of Mendelian Phenotypes: Discoveries, Challenges, and Opportunities. Am J Hum Genet 2015; 97: 199–215.

6. Evrony GD, Cordero DR, Shen J, et al. Integrated genome and transcriptome sequencing identifies a noncoding mutation in the genome replication factor *DONSON* as the cause of microcephaly-micromelia syndrome. Genome Res 2017; 27: 1323–1335.

7. Cummings BB, Marshall JL, Tukiainen T, et al. Improving genetic diagnosis in Mendelian disease with transcriptome sequencing. Sci Transl Med 2017; 9: eaal5209.

8. Kremer LS, Bader DM, Mertes C, et al. Genetic diagnosis of Mendelian disorders via RNA sequencing. Nat Commun 2017; 8: 15824.

9. LaMonte GM, Rocamora F, Marapana DS, et al. Pan-active imidazolopiperazine antimalarials target the Plasmodium falciparum intracellular secretory pathway. Nat Commun 2020 111 2020; 11: 1–15.

10. Lamonte G, Lim MYX, Wree M, et al. Mutations in the Plasmodium falciparum cyclic amine resistance locus (PfCARL) confer multidrug resistance. MBio; 7. Epub ahead of print 1 July 2016. DOI: 10.1128/mBio.00696-16.

11. Jonikas MC, Collins SR, Denic V, et al. Comprehensive Characterization of Genes Required for Protein Folding in the Endoplasmic Reticulum. Science (80-) 2009; 323: 1693–1697.

12. Zhang S, Xu C, Larrimore KE, et al. Slp1-Emp65: A Guardian Factor that Protects Folding Polypeptides from Promiscuous Degradation. Cell 2017; 171: 346–357.e12.

13. Baldwin BR, Keay S, Zhang C-O. Cloning and epitope mapping of a functional partial fusion receptor for human cytomegalovirus gH. J Gen Virol 2000; 81: 27–35.

14. Baldwin BR, Kleinberg M, Keay S. Molecular Cloning and Expression of Receptor Peptides That Block Human Cytomegalovirus/Cell Fusion. Biochem Biophys Res Commun 1996; 219: 668–673.

15. Symoens S, Barnes AM, Gistelinck C, et al. Genetic Defects in TAPT1 Disrupt Ciliogenesis and Cause a Complex Lethal Osteochondrodysplasia. Am J Hum Genet 2015; 97: 521–534.

16. Faghihi MA, Wahlestedt C. Regulatory roles of natural antisense transcripts Characteristics of antisense RNAs NIH Public Access. Nat Rev Mol Cell Biol 2009; 10: 637–643.

17. Pelechano V, Steinmetz LM. Gene regulation by antisense transcription. Nat Rev Genet 2013; 14: 880–893.

18. Lloret-Llinares M, Mapendano CK, Martlev LH, et al. Relationships between PROMPT and gene expression. RNA Biol 2016; 13: 6–14.

19. Seila AC, Core LJ, Lis JT, et al. Divergent transcription: a new feature of active promoters. Cell Cycle 2009; 8: 2557–2564.

20. Faghihi MA, Modarresi F, Khalil AM, et al. Expression of a noncoding RNA is elevated in Alzheimer’s disease and drives rapid feed-forward regulation of beta-secretase. Nat Med 2008; 14: 723–30.

21. Yu W, Gius D, Onyango P, et al. Epigenetic silencing of tumour suppressor gene p15 by its antisense RNA. Nature 2008; 451: 202–6.

22. Sigova AA, Mullen AC, Molinie B, et al. Divergent transcription of long noncoding RNA/mRNA gene pairs in embryonic stem cells. Proc Natl Acad Sci U S A 2013; 110: 2876–2881.

23. Hein MY, Hubner NC, Poser I, et al. A Human Interactome in Three Quantitative Dimensions Organized by Stoichiometries and Abundances. Cell 2015; 163: 712– 723.

24. Spaete RR, Mocarskit ES. Insertion and deletion mutagenesis of the human cytomegalovirus genome (herpesvirus/recombinant virus/expression vector/vaccine vector). 1987.

25. Arnold M, Bressin A, Jasnovidova O, et al. A BRD4-mediated elongation control point primes transcribing RNA polymerase II for 3’-processing and termination. Mol Cell 2021; 81: 3589–3603.e13.

26. Mayer A, Di Iulio J, Maleri S, et al. Native elongating transcript sequencing reveals human transcriptional activity at nucleotide resolution. Cell 2015; 161: 541–554.

27. Deng J, Li D, Mei H, et al. Novel deep intronic mutation in the coagulation factor XIII a chain gene leading to unexpected RNA splicing in a patient with factor XIII deficiency. BMC Med Genet 2020; 21: 9.

28. Naruto T, Okamoto N, Masuda K, et al. Deep intronic GPR143 mutation in a Japanese family with ocular albinism. Sci Rep 2015; 5: 11334.

29. Fusco C, Morlino S, Micale L, et al. Characterization of two novel intronic variants affecting splicing in FBN1-related disorders. Genes (Basel); 10. Epub ahead of print 1 June 2019. DOI: 10.3390/genes10060442.

30. Malekkou A, Sevastou I, Mavrikiou G, et al. A novel mutation deep within intron 7 of the GBA gene causes Gaucher disease. Mol Genet Genomic Med 2020; 8: e1090.

31. Patel N, Anand D, Monies D, et al. Novel phenotypes and loci identified through clinical genomics approaches to pediatric cataract HHS Public Access. Hum Genet 2017; 136: 205–225.

32. Worman HJ. Nuclear lamins and laminopathies. J Pathol 2012; 226: 316–325.

33. Frantz C, Stewart KM, Weaver VM. The extracellular matrix at a glance. J Cell Sci 2010; 123: 4195–4200.

34. Rozario T, DeSimone DW. The extracellular matrix in development and morphogenesis: a dynamic view. Dev Biol 2010; 341: 126–140.

35. Calleja-Agius J, Brincat M, Borg M. Skin connective tissue and ageing. Best Pract Res Clin Obstet Gynaecol 2013; 27: 727–740.

36. Forlino A, Marini JC. Osteogenesis imperfecta. Lancet 2016; 387: 1657–1671.

37. Guillemyn B, Nampoothiri S, Syx D, et al. Loss of TANGO1 Leads to Absence of Bone Mineralization. JBMR Plus 2021; 5: e10451.

38. Lekszas C, Foresti O, Raote I, et al. Biallelic TANGO1 mutations cause a novel syndromal disease due to hampered cellular collagen secretion. Elife; 9. Epub ahead of print 1 February 2020. DOI: 10.7554/ELIFE.51319.

39. Parvez MSA, Rahman MM, Morshed MN, et al. Novel insights into the structure and transport mechanisms of TAPT1. bioRxiv 2020; 2020.05.18.099887.

40. Maddirevula S, Alsahli S, Alhabeeb L, et al. Expanding the phenome and variome of skeletal dysplasia. Genet Med 2018; 20: 1609–1616.

41. Porntaveetus T, Theerapanon T, Srichomthong C, et al. Cole-Carpenter syndrome in a patient from Thailand. Am J Med Genet Part A 2018; 176: 1706–1710.

42. Ouyang L, Yang F. Cole-Carpenter syndrome-1 with a de novo heterozygous deletion in the P4HB gene in a Chinese girl. Med (United States); 96. Epub ahead of print 1 December 2017. DOI: 10.1097/MD.0000000000009504.

43. Balasubramanian M, Padidela R, Pollitt RC, et al. P4HB recurrent missense mutation causing Cole-Carpenter syndrome. J Med Genet 2018; 55: 158–165.

44. Li L, Zhao D, Zheng W, et al. A novel missense mutation in P4HB causes mild osteogenesis imperfecta. Biosci Rep; 39. Epub ahead of print 30 April 2019. DOI: 10.1042/BSR20182118.

45. Rauch F, Fahiminiya S, Majewski J, et al. Cole-carpenter syndrome is caused by a heterozygous missense mutation in P4HB. Am J Hum Genet 2015; 96: 425–431.

46. Howell GR, Shindo M, Murray S, et al. Mutation of a Ubiquitously Expressed Mouse Transmembrane Protein (*Tapt1*) Causes Specific Skeletal Homeotic Transformations. Genetics 2007; 175: 699–707.

47. Sohaskey ML, Jiang Y, Zhao JJ, et al. Osteopotentia regulates osteoblast maturation, bone formation, and skeletal integrity in mice. J Cell Biol 2010; 189: 511–525.

48. Friederichs JM, Gardner JM, Smoyer CJ, et al. Genetic Analysis of Mps3 SUN Domain Mutants in *Saccharomyces cerevisiae* Reveals an Interaction with the SUN-Like Protein Slp1. G3|Genes|Genomes|Genetics 2012; 2: 1703–1718.

49. Benham AM. The protein disulfide isomerase family: Key players in health and disease. Antioxidants and Redox Signaling 2012; 16: 781–789.

50. Annunen P, Helaakoski T, Myllyharju J, et al. Cloning of the human prolyl 4-hydroxylase α subunit isoform α(II) and characterization of the type II enzyme tetramer: The α(I) and α(II) subunits do not form a mixed α(I)α(II)β2 tetramer. J Biol Chem 1997; 272: 17342–17348.

51. Kukkola L, Hieta R, Kivirikko KI, et al. Identification and Characterization of a Third Human, Rat, and Mouse Collagen Prolyl 4-Hydroxylase Isoenzyme. J Biol Chem 2003; 278: 47685–47693.

52. Li H-J, Xue Y, Jia D-J, et al. POD1 Regulates Pollen Tube Guidance in Response to Micropylar Female Signaling and Acts in Early Embryo Patterning in *Arabidopsis*. Plant Cell 2011; 23: 3288–3302.

53. Maeda I, Kohara Y, Yamamoto M, et al. Large-scale analysis of gene function in Caenorhabditis elegans by high-throughput RNAi. Curr Biol 2001; 11: 171–176.

54. Kuhen KL, Chatterjee AK, Rottmann M, et al. KAF156 is an antimalarial clinical candidate with potential for use in prophylaxis, treatment, and prevention of disease transmission. Antimicrob Agents Chemother 2014; 58: 5060–5067.

55. Meister S, Plouffe DM, Kuhen KL, et al. Imaging of plasmodium liver stages to drive next-generation antimalarial drug discovery. Science (80-) 2011; 334: 1372–1377.

56. Dobin A, Davis CA, Schlesinger F, et al. STAR: ultrafast universal RNA-seq aligner. Bioinformatics 2013; 29: 15–21.

57. Frankish A, Diekhans M, Ferreira AM, et al. GENCODE reference annotation for the human and mouse genomes. Nucleic Acids Res 2019; 47: D766–D773.

58. Anders S, Pyl PT, Huber W. HTSeq-A Python framework to work with high-throughput sequencing data. Bioinformatics 2015; 31: 166–169.

59. Love MI, Huber W, Anders S. Moderated estimation of fold change and dispersion for RNA-seq data with DESeq2. Genome Biol 2014; 15: 550.

60. Shen S, Park JW, Lu ZX, et al. rMATS: Robust and flexible detection of differential alternative splicing from replicate RNA-Seq data. Proc Natl Acad Sci U S A 2014; 111: E5593–E5601.

61. Martin M. Cutadapt removes adapter sequences from high-throughput sequencing reads. EMBnet.journal 2011; 17: 10–12.

62. Gajos M, Jasnovidova O, Van Bommel A, et al. Conserved DNA sequence features underlie pervasive RNA polymerase pausing. Nucleic Acids Res 2021; 49: 4402–4420.

63. O’Leary NA, Wright MW, Brister JR, et al. Reference sequence (RefSeq) database at NCBI: current status, taxonomic expansion, and functional annotation. Nucleic Acids Res 2016; 44: D733–D745.

64. Kozomara A, Birgaoanu M, Griffiths-Jones S. miRBase: from microRNA sequences to function. Nucleic Acids Res 2019; 47: D155–D162.

65. Jurka J, Kapitonov V V., Pavlicek A, et al. Repbase Update, a database of eukaryotic repetitive elements. Cytogenet Genome Res 2005; 110: 462–467.

66. Li B, Dewey CN. RSEM: accurate transcript quantification from RNA-Seq data with or without a reference genome. BMC Bioinformatics; 12. Epub ahead of print 4 August 2011. DOI: 10.1186/1471-2105-12-323.

67. Davis CA, Hitz BC, Sloan CA, et al. The Encyclopedia of DNA elements (ENCODE): data portal update. Nucleic Acids Res 2018; 46: D794–D801.

68. Vangipuram M, Ting D, Kim S, et al. Skin punch biopsy explant culture for derivation of primary human fibroblasts. J Vis Exp. Epub ahead of print 2013. DOI: 10.3791/3779.

69. Nazari I, Tayara H, Chong KT. Branch point selection in RNA splicing using deep learning. IEEE Access. 2018 Dec 13;7:1800–7.

70. Paggi JM, Bejerano G. A sequence-based, deep learning model accurately predicts RNA splicing branchpoints. RnA. 2018 Dec 1;24(12):1647–58.

71. Zhang Q, Fan X, Wang Y, Sun MA, Shao J, Guo D. BPP: a sequence-based algorithm for branch point prediction. Bioinformatics. 2017 Oct 15;33(20):3166–72.

72. Jassal B, Matthews L, Viteri G, et al. The reactome pathway knowledgebase. Nucleic Acids Res 2020; 48: D498–D503.

