## Supplementary material for "Deciphering the consequence of deep intronic variants: a progeroid syndrome caused by a *TAPT1* mutation is revealed by combined RNA/SI-NET sequencing": Table S3

Table S3. List of PCR primers, LNA GapmeR sequences and qPCR primers

**Genotyping primer sequences**

| Gene | Primer | Sequence |
| --- | --- | --- |
| *TAPT1* | TAPT1-F1 | GGAAACCCCTGGCATAGACT |
|  | TAPT1-R1 | TGTGGACACGTGTAGCAAGA |
| *TAPT1* | TAPT1-F2 | CTGCAATGGCAGTCTTTTATTC |
|  | TAPT1-R2 | GGTTTCACAAGCAGGCAGAT |

**LNA GapmeR sequences**

| LNA GapmeR name | Sequence |
| --- | --- |
| *TAPT1-AS1* GapmeR 1 | GTTCCATCTCTTCTG |
| *TAPT1-AS1* GapmeR 2 | TCACTTACCTTCATGT |

**QPCR primer sequences**

| Gene | Primer | Sequence |
| --- | --- | --- |
| *TAPT1* | TAPT1-QPCR-F1 | AGACGCTGGGCTTCTACGA |
| *TAPT1* | TAPT1-QPCR-R1  TAPT1-QPCR-F2  TAPT1-QPCR-R2 | CCCTCTTGTTAGTTCAGCACTG  TGCTGAACTAACAAGAGGGTACT  TGAACACATACAAAAACGCATCC |
| *TAPT1-AS1* | TAPT1-AS1-QPCR-F1 | ATTTGGGCAAGAAGGAGCTT |
| *TAPT1-AS1* | TAPT1-AS1-QPCR-R1  TAPT1-AS1-QPCR-F2  TAPT1-AS1-QPCR-R2 | GGAGCTCCCTAAGGGCTAGA  CACCAGGCACTGCAATAAGA  CGACAGCATCGTCTCAAAGA |
| *RARRES2* | RARRES2-QPCR-F1 | AGAAACCCGAGTGCAAAGTCA |
|  | RARRES2-QPCR-R1 | AGAACTTGGGTCTCTATGGGG |
| *ZIC1* | ZIC1-QPCR-F1 | CACGCGGGACTTTCTGTTC |
|  | ZIC1-QPCR-R1 | TGCCCGTTGACCACGTTAG |
| *ZIC4* | ZIC4-QPCR-F1 | CCCTTCAGATGCGAGTTCGAG |
|  | ZIC4-QPCR-R1 | GTATGGCTTGTCGCTAGTGTG |
| *BIP* | BIP-QPCR-F1 | TGTTCAACCAATTATCAGCAAACTC |
|  | BIP-QPCR-R1 | TTCTGCTGTATCCTCTTCACCAGT |
| *ATF4* | ATF4-QPCR-F1 | GTTCTCCAGCGACAAGGCTA |
|  | ATF4-QPCR-R1 | ATCCTGCTTGCTGTTGTTGG |
| *CHOP* | CHOP-QPCR-F1 | AGAACCAGGAAACGGAAACAGA |
|  | CHOP-QPCR-R1 | TCTCCTTCATGCGCTGCTTT |
| *P4HB* | P4HB-QPCR-F1 | TCCTGGAGGGCAAAATCAAG |
|  | P4HB -QPCR-R1 | GGCATAGAACTCCACAAAGACG |
| *PDIA6* | PDIA6-QPCR-F1 | TTCTATGCTCCTTGGTGTGG |
|  | PDIA6-QPCR-R1 | GCCAGAACCTGATTGACTGTAG |
| *XBP1* (total) | XBP1 (total)-QPCR-F1 | TGGCCGGGTCTGCTGAGTCCG |
|  | XBP1 (total)-QPCR-R1 | ATCCATGGGGAGATGTTCTGG |
| *XBP1* (spliced) | XBP1 (spliced)-QPCR-F1 | CTGAGTCCGAATCAGGTGCAG |
|  | XBP1 (spliced)-QPCR-R1 | ATCCATGGGGAGATGTTCTGG |
| *GAPDH* | GAPDH-QPCR-F1 | CGACAGTCAGCCGCATCTT |
|  | GAPDH-QPCR-R1 | CCCCATGGTGTCTGAGCG |
